# 3D T2w fetal body MRI: automated organ volumetry, growth charts and population-averaged atlas

**DOI:** 10.1101/2023.05.31.23290751

**Authors:** Alena U. Uus, Megan Hall, Irina Grigorescu, Carla Avena Zampieri, Alexia Egloff Collado, Kelly Payette, Jacqueline Matthew, Vanessa Kyriakopoulou, Joseph V. Hajnal, Jana Hutter, Mary A. Rutherford, Maria Deprez, Lisa Story

## Abstract

Structural fetal body MRI provides true 3D information required for volumetry of fetal organs. However, current clinical and research practice primarily relies on manual slice-wise segmentation of raw T2-weighted stacks, which is time consuming, subject to inter- and intra-observer bias and affected by motion-corruption. Furthermore, there are no existing standard guidelines defining a universal approach to parcellation of fetal organs. This work produces the first parcellation protocol of the fetal body organs for motion-corrected 3D fetal body MRI. It includes 10 organ ROIs relevant to fetal quantitative volumetry studies. We also introduce the first population-averaged T2w MRI atlas of the fetal body. The protocol was used as a basis for training of a neural network for automated organ segmentation. It showed robust performance for different gestational ages. This solution minimises the need for manual editing and significantly reduces time. The general feasibility of the proposed pipeline was also assessed by analysis of organ growth charts created from automated parcellations of 91 normal control 3T MRI datasets that showed expected increase in volumetry during 22-38 weeks gestational age range. In addition, the results of comparison between 60 normal and 12 fetal growth restriction datasets revealed significant differences in organ volumes.

## Introduction

Fetal MRI provides complementary information to antenatal ultrasound that allows comprehensive assessment of fetal development and detection of abnormalities based on both visual inspection and quantitative analysis^1^. Structural MRI of the fetal body allows extraction of true 3D volumetric information via segmentation of individual organs and areas of abnormality for diagnosis and prognosis of outcomes^2^ or characterisation of normal fetal development^3^.

While dedicated acquisition protocols^4,5^ for structural fetal MRI such as T2 weighted single shot turbo spin echo (SSTSE) provide high in-plane image quality and contrast, the acquired stacks of slices are inherently corrupted by motion that leads to loss of spatial continuity in 3D. The recent developments in retrospective motion correction methods^6^ based on 3D deformable slice-to-volume registration (DSVR) for structural fetal body MRI^7,8^ allow reconstruction of high-resolution 3D isotropic images of the fetal body. The continuity of these images in 3D space provides superior visualisation (Davidson et al. 2021) as well as accurate 3D segmentation and volumetry of individual body organs including lungs^9^, thymus^10^ or heart^11^.

However, currently, there are no existing universally accepted standard protocols or guidelines for parcellation of body organs for fetal MRI. Different studies tend to rely on internal expertise^12^ with segmentation protocols adapted to specific research aims. Furthermore, in conventional clinical research practice, MRI-derived fetal body organ volumetry is primarily based on manual tracing in 2D planes^10,13,14^, which is time-consuming and sensitive to inter- and intra-observer bias. Volumetric information derived from 2D slice-wise segmentations might also vary between different stacks^15^.

Recently, there have been several studies that have used automated 3D parcellation for the fetal body organs^9,16,17^ based on atlas label propagation and deep learning for segmentation of the lungs and heart vessels. Yet, despite the successes in the application of deep learning for multi-label segmentation of the fetal brain^18^, to our knowledge, there has been no reported dedicated automated methods for simultaneous segmentation of multiple fetal body organs such as liver, spleen, thymus or kidneys.

### Contributions

In this work, we present the first multi-organs parcellation protocol for 3D motion-corrected T2w SSTSE MRI images of the fetal body. This protocol is used as a baseline for training of a deep learning pipeline for automated organ segmentation of 3D DSVR reconstructed body images based on semi-supervised training. The feasibility of the wider application of the pipeline is then tested based on segmentation of 91 normal fetal 3T T2w SSTSE MRI datasets for generation of volumetry growth charts of the normal fetal body organ development during 22-38 weeks GA range. In addition, we also use the proposed pipeline to compare organ volumes between the control cohort of normally developed fetuses and 12 cases of fetal growth restriction (FGR). We also present the first population-averaged 3D fetal MRI atlas of the normal fetal body.

## Methods

### Cohorts, datasets and pre-processing

The main fetal MRI cohort used in this work includes 138 T2w datasets from 21-38 weeks gestational age (GA) range acquired at St. Thomas’ Hospital, London. The datasets were collected prospectively as part of the Placental Imaging Project - PiP as approved by the Fulham Research Ethics Committee (REC 16/LO/1573) and Individualised Risk prediction of adverse neonatal outcome in pregnancies that deliver preterm using advanced MRI techniques and machine learning study as approved by the Fulham Research Ethics Committee (REC 21/SS/0082). All experiments were performed in accordance with relevant guidelines and regulations. Informed written consent was obtained from all participants.

The datasets (Fig. 1) were acquired on a 3T Philips Achieva MRI system using a 32-channel cardiac coil with a 2D T2w SSTSE sequence with TE=180ms. Each dataset includes 2-6 stacks covering the trunk ROI with voxel size=1.25x1.25x2.5mm, slice thickness=2.5mm, spacing=-1.5/2.5mm and 2-4 minutes acquisition time (depending on the GA and ROI coverage). The 3D reconstructions of the fetal trunk were performed using the original^7^ and automated^19^ DSVR pipelines in SVRTK (https://github.com/SVRTK/SVRTK). The output images have 0.8mm isotropic resolution and are reoriented to the standard radiological space.

**Figure 1.**
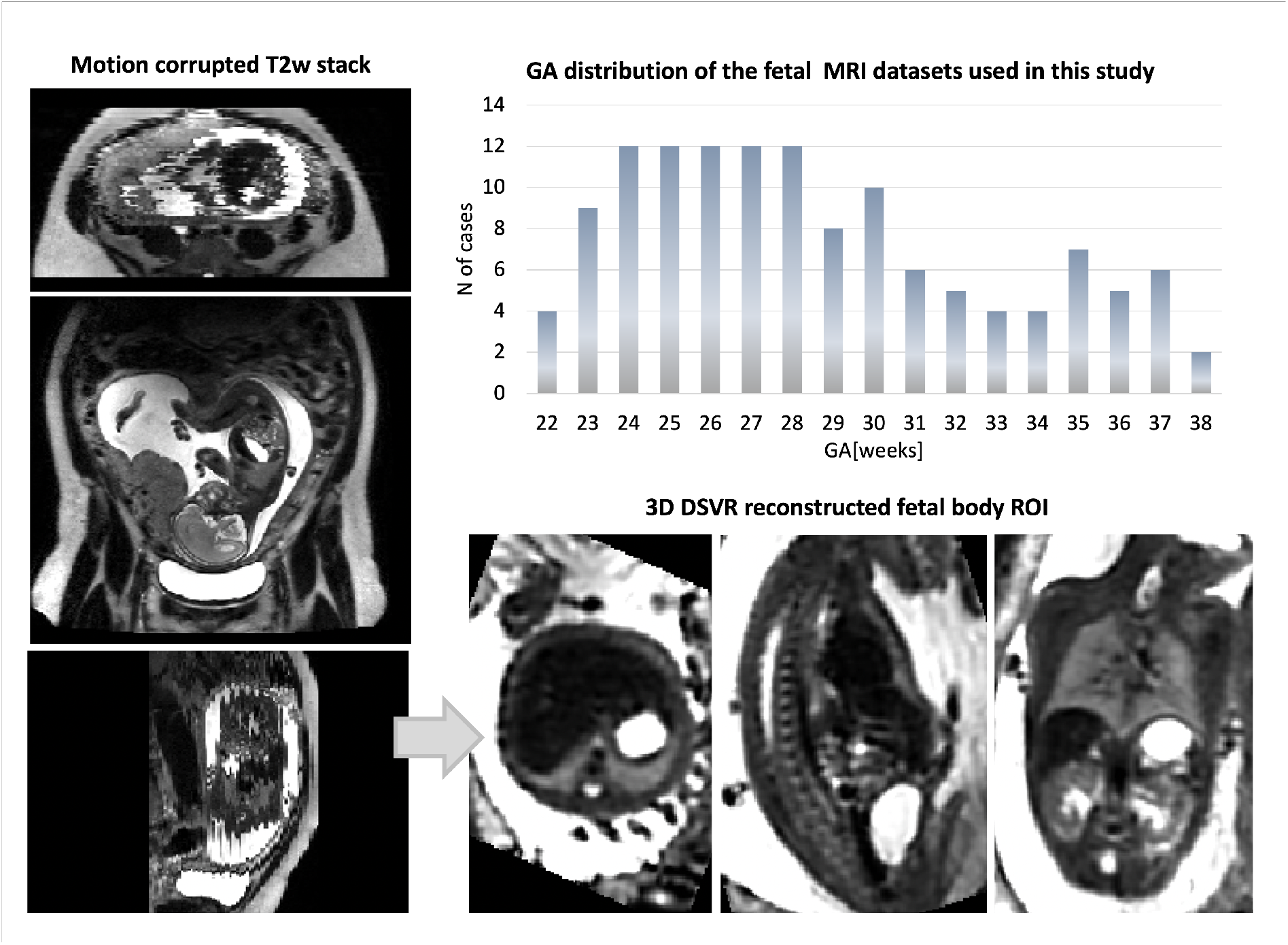
Fetal MRI 3T datasets used in this study: Gestational age distribution and an example of a 3D DSVR reconstructed fetal body.

The inclusion criteria were singleton pregnancy, no known structural abnormalities, acceptable DSVR reconstruction quality with visibility of all body organs for segmentation. Notably, while all included cases can be used for delineation and volumetry, the predominant part of the normal control cohort is suboptimal for body imaging due to the smaller number of available stacks with full trunk ROI coverage (because of the brain-only dedicated fetal T2w acquisition MRI protocol), which resulted in lower image quality.

The cohort used for training and testing of the deep learning pipeline includes 84 datasets of both control fetuses as well cases with reported placental anomalies or preterm birth risks factors. The 91 control datasets of normally developed fetuses that were used to generate organ growth charts do not have any diagnosed brain, body or placenta anomalies and were born at term. The additional 12 abnormal cases used for evaluation of feasibility of the segmentation method for quantitative studies were diagnosed with fetal growth restriction (estimated weight less than the 10th percentile).

In addition, we used 25 fetal MRI datasets from the Intelligent Fetal Imaging and Diagnosis project (REC 14/LO/1806) for preliminary training of the network. The datasets were acquired on 1.5T Philips Ingenia MRI system using 28-channel torso coil with TE=80ms and TE=180ms, acquisition resolution 1.25x1.25mm, slice thickness 2.5mm, -1.25mm gap and 9-11 stacks. The 3D images of the fetal body were reconstructed using^7^ to 0.8mm resolution and reoriented to the standard space.

### Definition of fetal body organ parcellation protocol

The protocol for segmentation was defined by a clinician (LS) with more than 15 years of experience in fetal MRI. The selected organs include: lungs, thymus, liver, spleen, stomach, gallbladder, renal parenchyma, renal pelvis, adrenal glands, and bladder. The selection criteria were based both on relevance to clinical quantitative volumetry studies and detection of anomalies as well as clear visibility in 3D DSVR fetal body images. At first, a set of example cases defining parcellation of individual organ ROIs were manually segmented slice-wise by clinicians (LS and MH) in ITK-SNAP^20^ in 3D DSVR images. The final version of the protocol is the output of the automated organ segmentation pipeline that corrected and smoothed irregularities and minor inconsistencies in continuity of the manual labels in 3D space.

### Population-averaged 3D atlas of the fetal body

In order to formalise the average parcellation model, we created a population-averaged fetal body atlas from 17 DSVR reconstructed images of normal subjects (25-28 weeks GA range). It was generated in MIRTK toolbox in 4 iterations using rigid, affine and non-rigid registration followed by averaging with Laplacian sharpening with 0.7mm isotropic resolution. The atlas was segmented by the network followed by additional manual refinement of local features. The atlas and the proposed parcellation protocol are publicly available online at KCL CDB fetal MRI body atlas repository (https://gin.g-node.org/kcl_cdb/fetal_body_mri_atlas).

### Automated segmentation of fetal body organs

The pipeline for automated deep learning multi-label 3D segmentation of the fetal body organs in 3D DSVR MRI images is shown in Fig. 2. At first, the trunk region is globally localised using a single-label network to exclude any surrounding background. Next, the resulting masked and cropped trunk image is used for 3D segmentation of 10 organ ROIs based on the defined protocol. Finally, the labels are transformed to the original image space.

**Figure 2.**
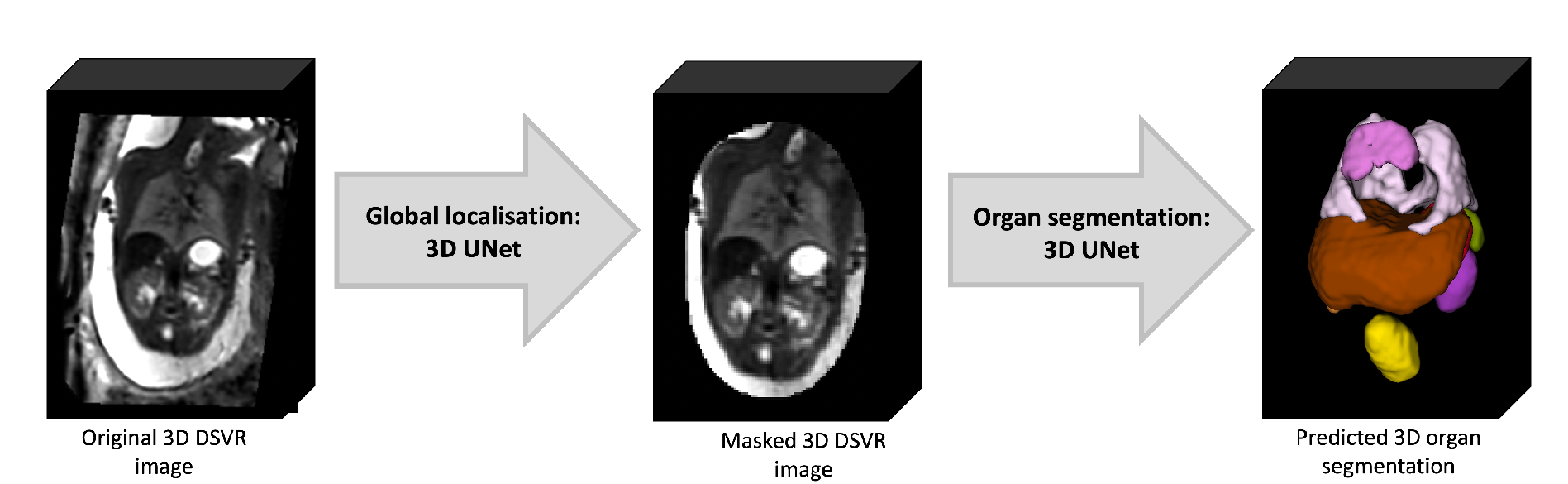
Proposed 3D fetal body organ segmentation pipeline.

The deep learning pipeline was implemented based on Pytorch MONAI^21^ framework using the classical 3D UNet^22^ architecture with five encoder-decoder blocks (output channels 32, 64, 128, 256 and 512), convolution and upsampling kernel size of 3, ReLU activation, dropout ratio of 0.5, batch normalisation, and a batch size of 12. We employ AdamW optimiser with a linearly decaying learning rate, initialised at 1x10-3, default beta parameters and weight decay 1x10-5. The preprocessing steps included masking to the global body ROI (pretrained 3D UNet from^19^) and resampling with padding to a 128x128x128 grid. We used standard MONAI augmentation including bias field and affine rotations (+/- 30 degrees).

The training was performed in two stages. At first, a set of 40 mixed 1.5T and 3T DSVR images from other studies at our department were manually segmented and refined (by LS, MH and AU) in ITK-SNAP while the lungs and thymus labels propagated from an in-house fetal thorax atlas^23^ using non-rigid registration in MIRTK and manually refined. These segmentations were used to pretrain the preliminary version of the network for 10000 iterations with MONAI-based augmentation (bias field, gaussian noise and rotations) and additional MIRTK histogram matching. Next, the pipeline was employed to segment the final set of 78 3T DSVR images that were manually refined. The final version of the network was trained using 70 training and 8 validation datasets for 50000 iterations.

The performance was tested on 5 datasets from 23-38 weeks vs. manual labels in terms of the organ detection status (qualitative visual assessment: correct=100%, partial=50%, failed=0%) and Dice. The manual labels segmented 2D slice-wise by MH using different planes for different organs and further refined by AU using 3D bush to correct major discontinuities in 3D.

### Implementation details

The MONAI-based source code and trained models are publicly available in 3D Fetal MRI GitHub repository (https://github.com/3DFetalMRI/fetal-body-organ-segmentation). The full segmentation pipeline along with the automated 3D reconstruction is publicly available as a standalone docker at SVRTK Segmentation repository (https://hub.docker.com/r/fetalsvrtk/segmentation; tag: *t2_body_organs*).

### Generation of normal body organ volumetry growth charts

We used the proposed deep learning pipeline to segment 91 3D DSVR fetal body images of 82 control fetuses from 22-38 weeks GA range. All segmentations were reviewed and manually refined, if required. The organ label volumes were used to generate normal organ development growth charts with mean, 5th and 95th centiles (Royston and Wright 1998) and assess the global volumetry trends.

### Comparison of normal and abnormal cohorts

In addition, in order to evaluate the feasibility of using the proposed pipeline for quantitative clinical studies, we used the automated segmentations for comparison of organ volumetry between the normal control (60) and FGR (12) datasets for 21-31 weeks GA range based on ANCOVA performed in Python statsmodels module.

## Results

### Proposed fetal body organ parcellation protocol

Fig. 3 shows the proposed 3D body organ parcellation protocol at different GAs: 22, 29 and 36 weeks. All organs have smooth boundaries defined based on both signal intensities and organ anatomy. The major cardiac and pulmonary vessels and trachea were excluded from the lung and thymus labels based on their visibility and homogeneity of the organ tissue. The liver ROI has inhomogeneous texture and includes the vasculature (e.g., the main portal vein). The kidneys are separated into parenchyma and pelvis regions. The adrenal glands are located above the kidneys. The spleen has homogeneous intensity and separated from the bowel. The boundaries of the fluid-filled ROIs (stomach, gallbladder, kidney pelvis and bladder) were defined based on the high signal intensity interface and the partial volume effect. While the tissue contrast of the organs (especially the lungs) visibly increases with the GA, visual inspection of the 3D models (Fig.3) demonstrates that relative positioning, shape and proportions between the organs remain similar at early and late GA.

**Figure 3.**
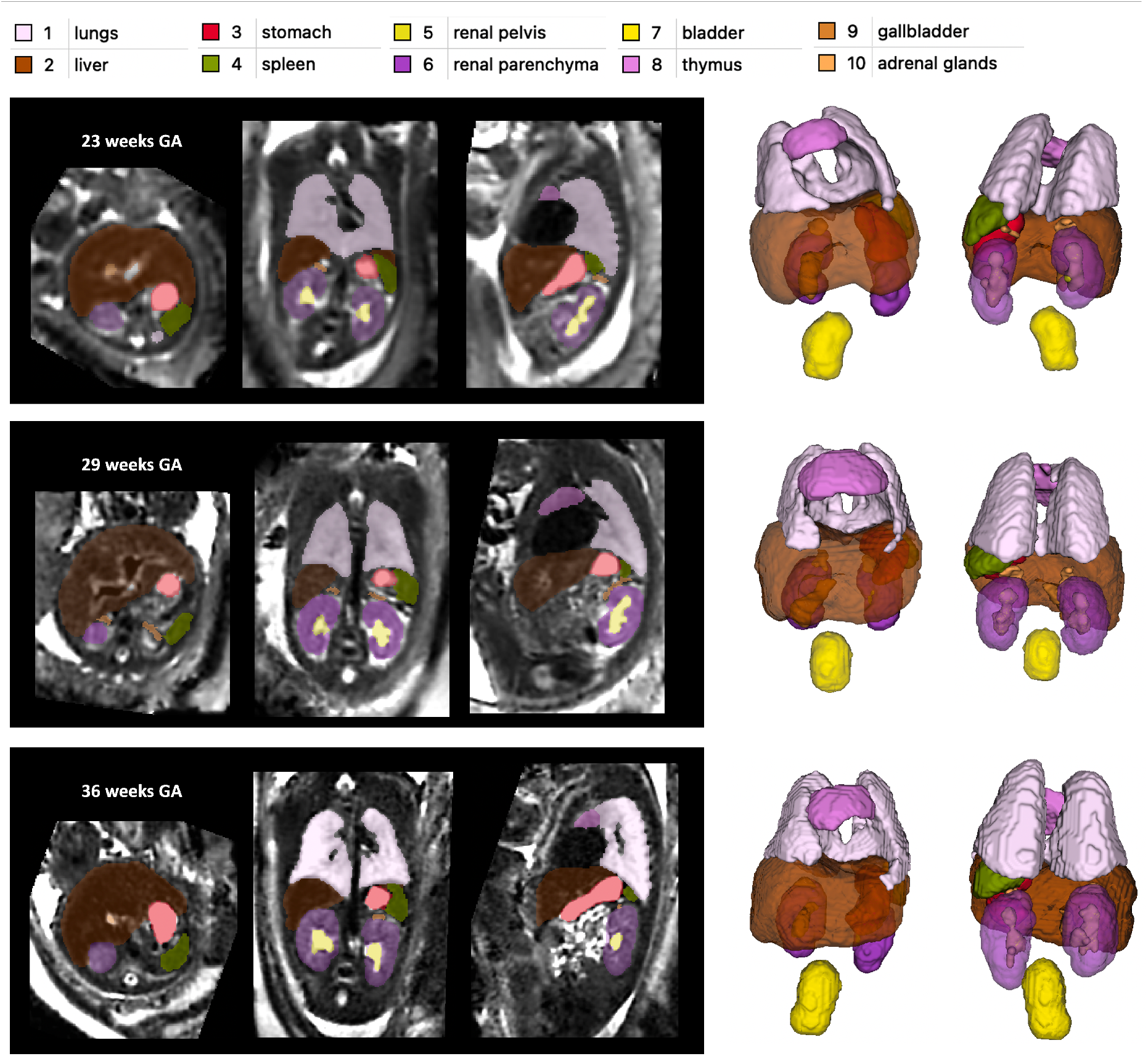
Proposed body organ parcellation protocol for 3D T2w fetal MRI. Examples at 23, 29 and 36 weeks GA.

### Population-averaged 3D atlas of the fetal body

The created average 3D body MRI atlas is shown in Fig. 4. It has continuous and well defined organ features. The atlas was inspected by LS and the anatomy was confirmed to be correct and corresponding to normal fetal development. The organ segmentations have clear boundaries at tissue interfaces and 3D models have similar appearance to individual subjects.

**Figure 4.**
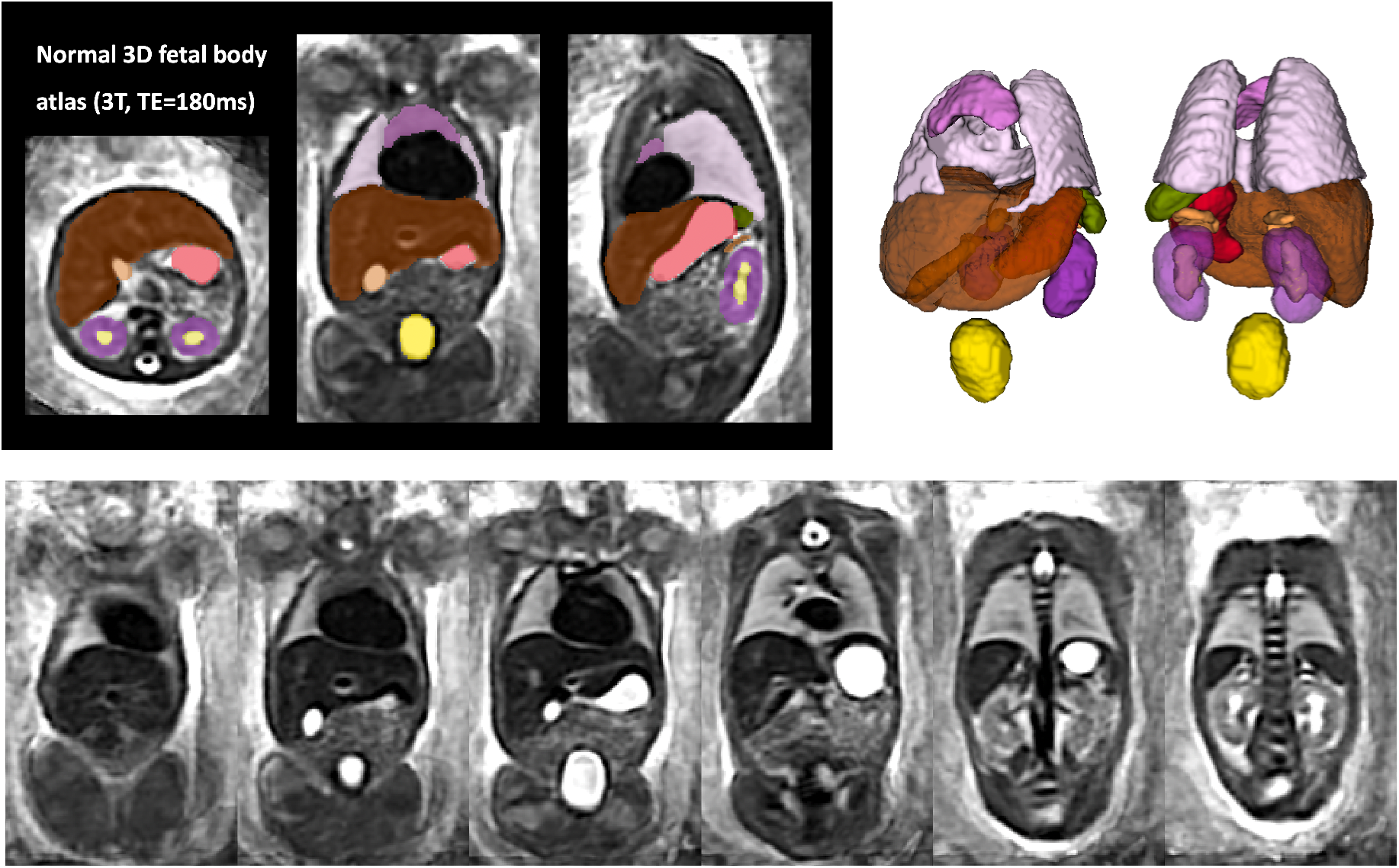
Created 3D average T2w fetal body atlas and the corresponding organ segmentation.

### Automated segmentation of fetal body organs

The results of the assessment of the network performance on 5 test cases from different GA groups are presented in Fig. 5A. The UNet was able to reliably detect all organs in all test subjects (94% due to partial errors for adrenal glands and kidneys).

**Figure 5.**
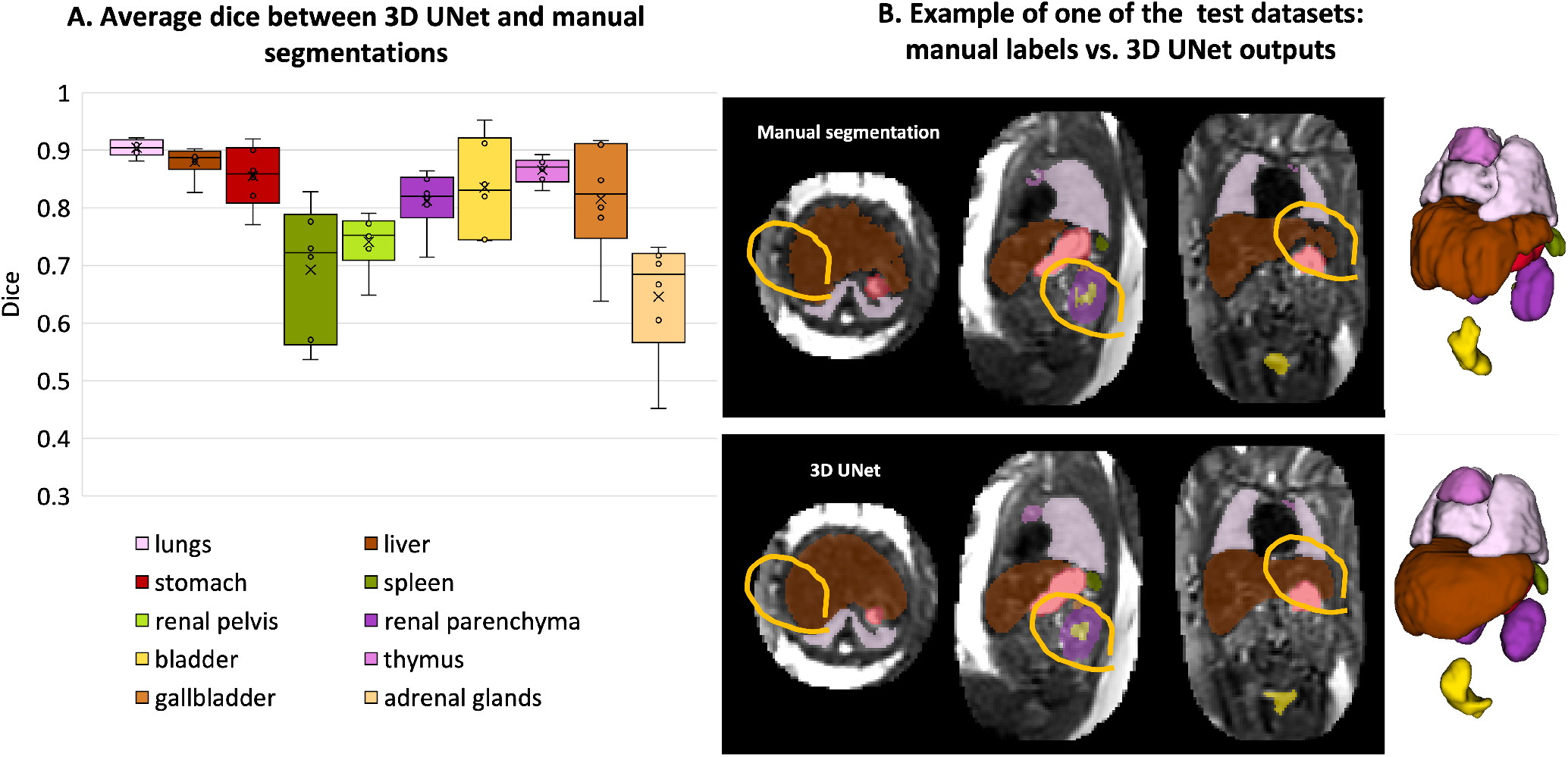
Results of testing of the proposed automated body organ parcellation protocol for 5 datasets: average Dice values for individual organs (A); visual comparison between manual and 3D UNet labels (B) (the red circles highlight the areas with differences in labels).

Notably, manual labels are prone to inconsistencies and discontinuities in 3D and cannot be considered as an absolute ground truth (Fig. 5B). This, along with the smoother boundaries produced by the network, is reflected in the moderately high Dice values for the medium size organ ROIs (thymus, spleen, gallbladder, renal pelvis). The Dice values for lungs, liver, stomach, bladder and renal parenchyma are high due to the large size and high contrast. This is in agreement with the fact that larger organs with smooth boundaries will naturally have larger Dice scores than small organs or organs with complex boundaries. The low values for adrenal glands are due to the small size as well as the partial detection and inconsistencies of manual segmentations in the training datasets that propagated to the network predictions. This indicates that, in future, additional refinement of the training labels for adrenal glands, thymus, spleen and renal pelvis will be required for further improvement of the network performance.

While these proof-of-concept results confirm the general suitability of deep learning for fetal organ volumetry, this also highlights that automated segmentation outputs should always be reviewed and fine-edited, if required.

### Normal body organ volumetry growth charts

The volumetry growth charts generated from automated organ segmentations of 91 normal control MRI datasets are given in Fig. 6. In less than 25% of cases, minor refinements were required predominately for the liver and kidney ROIs. This was primarily due to suboptimal image quality of the control datasets and lower contrast in the abdomen region and similarity to the bowel intensity. This, however, did not produce a significant difference in the results on the global trends with <5% variation for organ volumes in individual subjects. Notably, the time required for correction of the automated labels is on average <5 minutes per case while the conventional fully manual segmentation approach requires 2-3 hours per case.

**Figure 6.**
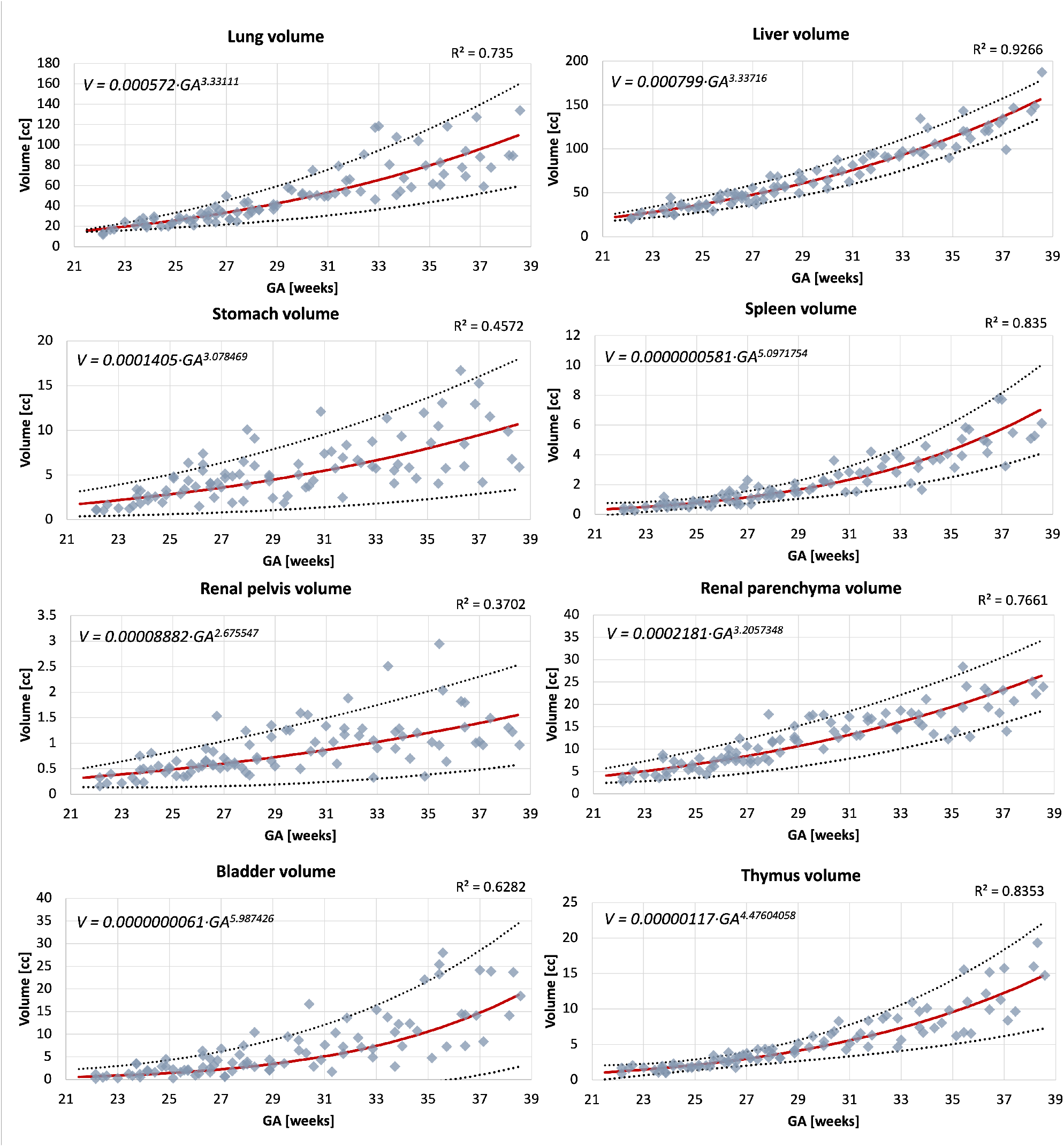
Volumetry growth charts of normal fetal body organ development created from the automated segmentations (the main 8 ROIs) of 91 normal control 3D motion-corrected 3T T2w fetal MRI datasets with 5th, mean, and 95th centiles.

The growth charts show the expected increase in the volume for all solid tissue organs. The similar increasing trends are also present for the fluid-filled organs, even though their volumes are not directly defined by GA. Notably, there is a pronounced wider individual variation with age of the lung volumes in the third trimester which is also visible in the earlier reported nomograms for 2D fetal MRI^24^ and ultrasound^25^. The volumes for the rest of the solid organs appear to be more consistent. The lung, liver and spleen volume trends are within the ranges of the earlier reported 3D ultrasound-derived nomograms^26,27^.

### Comparison of normal and abnormal cohorts

The results of comparison of 12 FGR with 60 normal control cases are presented in Fig. 7. Even taking into account the small number of available FGR cases, there is a significant difference in the volumes of lungs (p<0.05), liver (p<0.01) and renal parenchyma (p<0.01), which is in agreement with the expected smaller fetal size in FGR cohort^28,29^.

**Figure 7.**
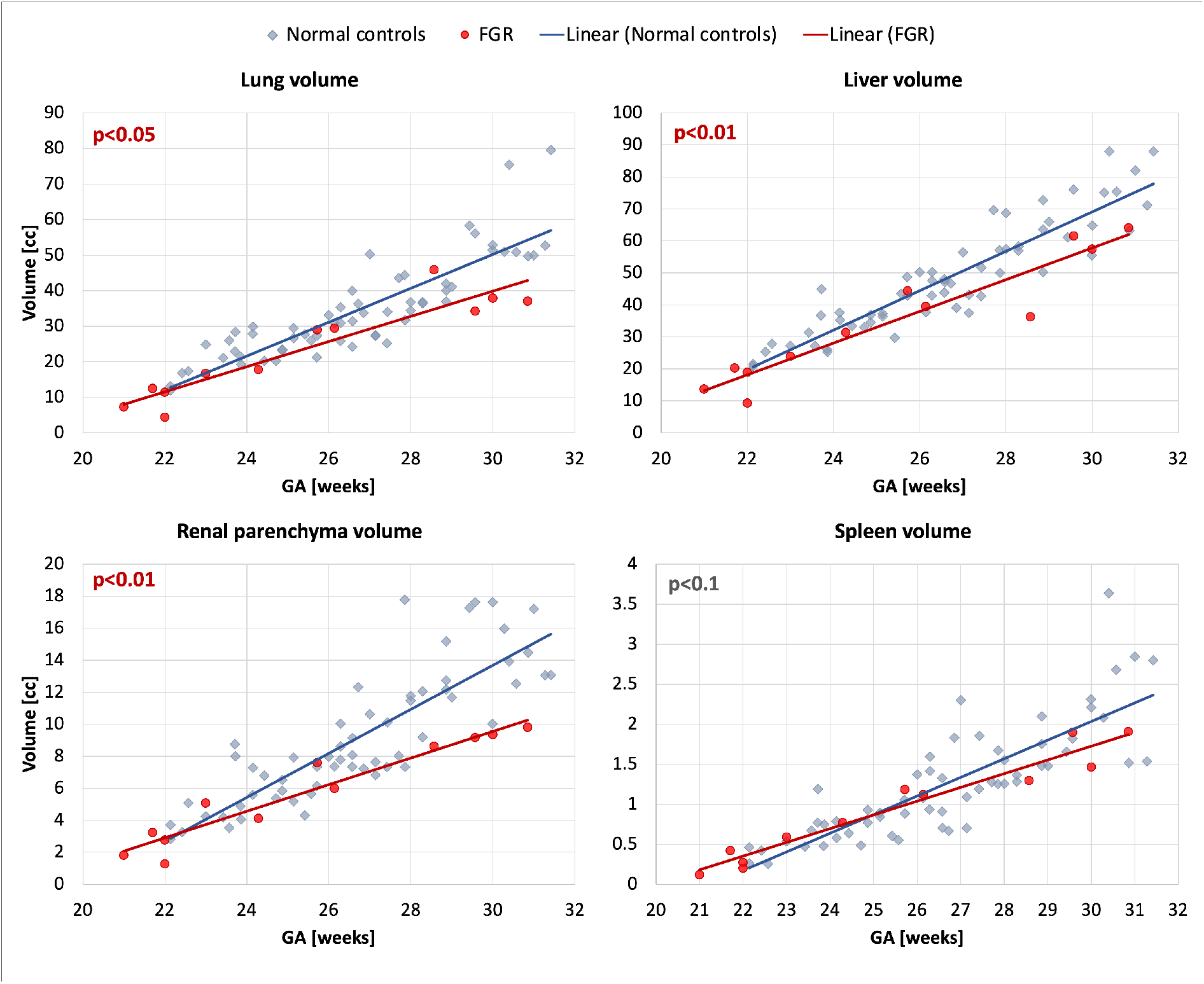
Comparison of FGR (12) and normal control (60) cohorts at 21-31 weeks GA range based on automated segmentation of 3D motion-corrected 3T T2w fetal MRI datasets: lung, liver, renal parenchyma and spleen volumes.

## Discussion

Fetal body organ volumetry is increasingly being used for quantitative clinical fetal MRI studies. However, it has been conventionally based on 2D slice-wise segmentation of motion-corrupted stacks, which is time consuming and subject to errors and inter-observer variabilities. Statistically accurate normal organ volume ranges are essential for differentiation of normality from pathology. In fetal imaging this has been persistently difficult to achieve: in ultrasound because of restricted field of view, difficulty establishing anatomical landmarks and variation in post-processing techniques^30^; and in MRI because of acquisition difficulties and limited post-processing techniques. Previous work has demonstrated wide variation in suggested normal ranges for fetal lung volumes, likely secondary to differing acquisition protocols and inability to use consistent slices for volume estimation owing to fetal motion^31^. Furthermore, the lack of standardised guidelines for organ parcellation limits direct comparison between different studies.

In this work, we defined the first 3D parcellation protocol for motion-corrected T2w SSTSE fetal body MRI with 10 organs ROIs relevant to quantitative volumetry studies and with good visibility in 3D DSVR images. Definition of the organ labels was based on both signal intensities and anatomical interfaces. This protocol was then used as a basis for creating manual labels for training a deep learning pipeline for 3D automated organ segmentation. Quantitative evaluation based on comparison with manual labels demonstrated robust performance for different GAs. The main challenges for the proposed pipeline were separation between the bowel and the liver, spleen and renal parenchyma ROIs due to similar intensity range and segmentation of the adrenal glands due to the small size and limited visibility.

Next, we used the proposed pipeline to segment a cohort of 91 normal control datasets and 12 FGR cases. Only minimal manual refinement was required in <25% of cases. This is a significant improvement in terms of time (<5 minutes per case) vs. the conventional manual segmentation approach (1.5-3 hours per case) as well as continuity of organ ROIs. Analysis of the generated normal growth charts demonstrated the expected increase with GA with the volume ranges being similar to the reported values from 3D ultrasound measurements. Comparison between the normal and FGR cohorts also showed a significant difference in major solid organs even despite the small number of available FGR cases.

These preliminary results confirmed that the proposed automated 3D organ segmentation pipeline is potentially suitable for analysis of large fetal MRI studies. The use of consistent acquisition protocols and 3D isotropic DSVR-reconstructed images also reduces the impact of fetal motion on segmentation of continuous organ ROIs which inherently improves accuracy of volumetric results in comparison to conventional 2D-based approach. Yet, the testing of the pipeline also highlighted the challenges related to segmentation of smaller structures and the need for extension of the protocol with more organ ROIs (e.g., bowel) along with optimisation for pathologies. Furthermore, all datasets have the same acquisition protocol (3T, TE=180ms) and the organ appearance in terms of signal intensity and contrast might vary for different sequence parameters and field strengths.

This is a first step towards standardisation of automated 3D MRI fetal body organ volumetry for quantitative analysis. However, translation of this solution for any quantitative studies of datasets from different acquisition protocols will require further detailed analysis of differences in image-derived features and implementation of a harmonisation solution. Furthermore, the organ volumetry nomograms should also include analysis of the impact of maternal parameters, sex and ethnicity as well as normalisation with respect to the whole body volume. After establishment of normality, this technique can be utilised to define anomalies antenatally, for example pulmonary hypoplasia which remains difficult to predict by ultrasound, particularly when secondary to oligohydramnios^32^. Additionally, conditions such as pre-eclampsia, fetal growth restriction and chorioamnionitis are all thought to alter volumetry of organs variably. Our future work will also include extension of the list of organs, further refinement of segmentations at tissue interfaces and for small ROIs, separation into left and right regions and optimisation of the protocol for abnormal cases as well as whole body segmentation.

## Conclusions

In this work, we introduced the first parcellation protocol and automated pipeline for multi-organ segmentation of motion corrected T2w 3D fetal body MRI. It was used to generate volumetry growth charts of normal fetal organ development during 22-38 weeks GA range. The results demonstrated robust performance of the pipeline for different gestational ages with only minor manual refinements required in less than 25% of cases that did not produce significant differences in volumetry trends. Furthermore, the automated segmentation-based analysis detected the expected difference between FGR and normal cohorts. This suggests the potential feasibility of using automated segmentation of large-scale quantitative volumetry studies that would significantly reduce the need for extensive manual input and minimise inter- and intra-observer variability.

Our future work will focus on extension of the list of organs, further refinement at tissue interfaces, separation into left and right regions as well as optimisation of the protocol for abnormal cases and harmonisation of the segmentation pipeline for various acquisition protocols.

## Acknowledgements

We thank everyone who was involved in acquisition and analysis of the datasets at the Department of Perinatal Imaging and Health at Kings College London and St Thomas’ Hospital. We thank all participants and their families.

This work was supported by NIHR Advanced Fellowship awarded to Lisa Story [NIHR30166], by the Wellcome Trust, Sir Henry Wellcome Fellowship to Jana Hutter, [201374/Z/16/Z], by the UKRI, FLF to Jana Hutter [MR/T018119/1], MRC grant [MR/W019469/1], the Wellcome Trust and EPSRC IEH award [102431] for the iFIND project [WT 220160/Z/20/Z], the Wellcome/ EPSRC Centre for Medical Engineering at King’s College London [WT 203148/Z/16/Z], MRC Confidence in concept [MC_PC_19041], the NIH Human Placenta Project grant [1U01HD087202-01], the NIHR Clinical Research Facility (CRF) at Guy’s and St Thomas’ and by the National Institute for Health Research Biomedical Research Centre based at Guy’s and St Thomas’ NHS Foundation Trust and King’s College London.

The views expressed are those of the authors and not necessarily those of the NHS, the NIHR or the Department of Health.

## Author contributions

A.U. prepared the manuscript, processed the datasets, trained the networks, created the atlas and produced the graphs. M.H. produced manual segmentations, analysed the results and prepared the clinical descriptions for the manuscript. I.G., K.P. participated in analysis of the results. C.A.Z., A.E.C., J.M., V.K. contributed to formalisation of the fetal body segmentation protocol. J.V.H. provided acquisition techniques and fetal MRI datasets. J.H. provided acquisition techniques and fetal MRI datasets and participated in analysis of the results. M.R. provided datasets and participated in analysis of the results. M.D. participated in analysis of the results. L.S. provided datasets, produced manual segmentations, formalised the segmentation protocol, participated in analysis of the results and supervised the research. All authors reviewed the manuscript.

## Data availability

The individual fetal MRI datasets used for this study are not publicly available due to ethics regulations. The created average normal 3D T2w atlas is publicly available online at KCL CDB fetal body MRI atlas repository: https://gin.g-node.org/kcl_cdb/fetal_body_mri_atlas.

